# To ban or not to ban social media for children? Beliefs and influencing factors among Greek parents

**DOI:** 10.64898/2026.06.26.26356645

**Authors:** A. Katsiroumpa, I. Moisoglou, P. Gallos, O. Galani, M. Tsiachri, P. Peleka, A. Triantafillaki, A. Kolisiati, P. Galanis

## Abstract

**OBJECTIVE:** To examine parents’ perceptions regarding the introduction of a social media ban for children and to identify factors associated with these attitudes.

**METHOD:** A cross-sectional study was carried out in Greece in April 2026. Potential predictors of parents’ views on a social media ban included (a) sociodemographic variables (such as gender, age, educational attainment, and financial status), (b) social media usage patterns (number of accounts, daily usage duration, and posting frequency), and (c) level of political engagement (how often participants follow political news and discuss political issues). Outcome variables comprised parents’ agreement with the ban, level of awareness about its implementation, perceived necessity for additional measures, confidence in the ban’s effectiveness, perceived effects on children’s lives, and parents’ familiarity with digital parental control tools.

**RESULTS:** Overall, 68.0% of parents supported implementing a social media ban for children under 15. A large majority (91.8%) expressed the need for more governmental information regarding the ban. Additionally, 89.3% believed that further measures beyond the ban are required to effectively address the issue. Suggested measures included digital literacy courses in schools (86.1%), active parental involvement in digital literacy (74.6%), prohibition of inappropriate content (77.9%), reasonable parental limits on social media use (73.8%), and restriction of addictive platform features (73.0%). Older parents demonstrated greater confidence in the effectiveness of the ban. Furthermore, age, financial status, number of social media accounts, and time spent online were positively associated with perceived impacts of the ban. Younger age was linked to greater parental familiarity with digital control tools, while having more social media accounts was also positively associated with such familiarity.

**CONCLUSIONS:** There is a clear need for comprehensive, evidence-based policy approaches that combine regulation, education, and shared responsibility among stakeholders. Policymakers should leverage existing public support for child protection while investing in digital literacy initiatives, empowering parents, and strengthening regulatory oversight of social media platforms to achieve long-term and equitable results.

## Introduction

International organizations and policymakers have increasingly begun to re-evaluate current digital governance frameworks, with growing emphasis on measures such as age-based restrictions, stronger platform accountability, and in some cases, the implementation of social media bans for children. For instance, the European Union has introduced a broad range of legislative and policy initiatives aimed at protecting children in digital environments while also enabling their meaningful participation online. These initiatives include regulations covering digital services, audiovisual media, and data protection, alongside practical tools such as helplines and reporting mechanisms for harmful or illegal content. Furthermore, the European Union has established a digital identity framework designed to facilitate secure and reliable age verification, thereby helping to limit children’s exposure to content that is inappropriate for their age.

Furthermore, governments at both national and regional levels have taken steps to strengthen child protection in digital environments, with many countries implementing age limits and verification systems. While some jurisdictions have introduced bans on smartphone use within schools, others have prioritized preventive approaches, including awareness campaigns and educational initiatives aimed at enhancing digital literacy among both children and parents. At the same time, civil society organizations and international bodies play an important role in promoting online child safety through research, public outreach, and specialized support services. Globally, the adoption of school smartphone bans has expanded significantly, with 58% of countries now enforcing such measures. Evidence from a recent meta-analysis suggests that smartphone bans in schools have a statistically significant, though relatively modest, overall impact. Their effects appear more pronounced in improving social wellbeing than in enhancing academic performance, with reductions observed in issues such as bullying.^1^

Australia is the first country to implement a nationwide restriction on social media access for individuals under the age of 16. The law came into effect on 10 December 2025 and does not allow parental consent as an exception to this age requirement. As a result, from that date onward, individuals younger than 16 are prohibited from creating or maintaining accounts on major social media platforms, including Facebook, TikTok, Instagram, YouTube, Snapchat, and X, among others. While the measure has been framed as a precautionary response to perceived shortcomings of platform self-regulation and parental mediation, it has also generated substantial public and academic debate, particularly regarding feasibility, circumvention, and potential implications for children’s rights and social participation.^2–6^

In this rapidly changing landscape, Greece’s planned social media ban for children under the age of 15, set to take effect on January 1, 2027, reflects a broader international policy shift. This development highlights the need to examine parents’ attitudes and their underlying determinants in order to evaluate the social legitimacy and practical feasibility of such measures. Since the literature has already showed the negative association between problematic social media use and mental health in studies in Greece^7–13^ there is an urgent need to further explore social media issues such as the opinion towards social media ban for children. Accordingly, the present study aimed to investigate parents’ opinion in Greece regarding social media ban for children and to identify the factors shaping parents’ views on this policy.

More specifically, the study explored the role of sociodemographic characteristics, patterns of social media use, and levels of political engagement as possible predictors of attitudes toward this measure. To the best of our knowledge, this represents the first study addressing this topic worldwide. By analyzing parents’ perspectives on restricting children’s access to social media, the study seeks to contribute to the development of informed public health strategies and evidence-based policy decisions aimed at enhancing child protection in the digital environment. Recently, we investigated general population opinion toward social media ban for children and its influencing factors.^14^ We further expand our research by examining in this study the beliefs of parents with children aged 8-17 years towards social media ban for children. Moreover, we investigated sociodemographic factors, social media use and political interest as potential predictors of parents’ beliefs towards social media ban.

## Methods

### Study design

On April 8, 2026, the Greek government declared that, effective January 1, 2027, children under the age of 15 will be barred from accessing social media. This restriction targets platforms that enable users to create personal accounts, engage with others, and share publicly accessible content, including posts, stories, videos, comments, likes, and follower interactions. Applications such as Facebook, Instagram, TikTok, and Snapchat fall under this definition. As a result, social media services operating within Greece will be obligated to introduce age-verification systems for all users by January 1, 2027. These systems may involve methods such as verification through official documents (e.g., identity cards or passports) or biometric technologies (e.g., facial recognition or age estimation). However, the exact procedures for age verification have not yet been finalized, as efforts continue at the European Union level to establish harmonized regulations. Responsibility for enforcing compliance will lie with the platforms themselves. Nonetheless, the prohibition of social media use by individuals younger than 15 years will take effect in Greece from January 1, 2027.

Within this framework, a cross-sectional study was carried out in Greece two weeks following the government’s announcement regarding the restriction on social media use among children under 15. Data collection took place in April 2026 through an online survey. The questionnaire, designed using Google Forms, was disseminated via social media platforms, including Facebook, Instagram, TikTok, and LinkedIn, yielding a convenience sample. Parents were eligible if they: (a) were aged 18 years or older, (b) had children aged 8-17 years, (c) provided informed consent, (d) were proficient in the Greek language, given that the study was conducted in Greece, (e) had internet access, and (f) maintained an active account on at least one social media platform. The study was conducted in accordance with the Strengthening the Reporting of Observational Studies in Epidemiology (STROBE) guidelines.^15^

## Measurements

### Sociodemographic characteristics

We assessed several sociodemographic variables as potential predictors of parents’ beliefs toward banning social media use among children. These included: (1) sex (male or female), (2) age (treated as a continuous variable), (3) level of education (categorized as some high school or less, high school diploma, or college degree), and (4) financial status (measured using a self-reported scale ranging from 0 [very poor] to 10 [very good]).

### Social media use

Social media use was assessed through the following measures: (1) the number of social media accounts held by parents (including platforms such as Facebook, Instagram, TikTok, YouTube, X [formerly Twitter], LinkedIn, and Snapchat), (2) the amount of time spent on social media each day (treated as a continuous variable), and (3) the frequency of posting on social media, categorized as never, approximately once per month, once every two weeks, once per week, two to four times per week, or five to seven times per week.

### Political engagement

Political engagement was assessed using two items. The first item evaluated the extent to which parents follow political news, with responses recorded on a five-point Likert scale ranging from 1 (“not closely at all”) to 5 (“very closely”). The second item measured the frequency with which parents discuss politics with others, also using a five-point Likert scale ranging from 1 (“not at all”) to 5 (“very often”). The scores from both items were summed up to produce an overall index ranging from 2 to 10. This composite measure represented the variable “political engagement” with higher scores indicating greater levels of engagement.

### Opinion on social media ban for children

Several questions were used to measure parents’ opinion on social media ban for children, such as “In your opinion, at what age is it appropriate for a child to have a personal social media account?”, “In your opinion, who should be responsible for setting limits on children’s use of social media?”, and “To what extent do you agree with the fact that the problematic social media use among children constitutes an important public health concern?”.

Furthermore, to assess parents’ confidence in the effectiveness of the social media ban, we used three items: “Children will be able to find ways to create accounts on platforms to which the social media ban will be applied”, “Children will create accounts on platforms that will be free and, therefore, less regulated, and “Social media platforms will fully comply with the legislation regarding the social media ban”. Responses to these three items were recorded using a five-point Likert scale ranging from 1 (“strongly disagree”) to 5 (“strongly agree”). The first two items were reverse-coded to ensure that higher scores uniformly reflected greater confidence in the effectiveness of the social media ban across all items. Subsequently, the item scores were summed to produce a total score ranging from 3 to 15. This aggregate score represented the variable “confidence in the effectiveness of the social media ban” with higher values indicating stronger confidence in its effectiveness.

To assess parents’ views regarding the impact of the social media ban on children’s lives, the following items were used: (1) Social media ban will improve children’s mental health, (2) Social media ban will improve children’s sleep quality, and (3) Social media ban will improve children’s school performance. Responses to these three items were collected using a five-point Likert scale ranging from 1 (“strongly disagree”) to 5 (“strongly agree”). The scores from the three items were then summed up to produce a total ranging from 3 to 15. This aggregate score represented the variable “impact of the social media ban on children’s lives,” with higher scores indicating a more positive perceived impact of the ban on children’s lives.

In addition, we examined whether parents perceived social media ban as more or less effective compared with alternative measures, such as restriction of the addictive features of social media platforms, prohibition of platforms from displaying inappropriate content, and integration of the digital literacy courses into the school curriculum.

Finally, parents were asked to indicate their level of familiarity with, and ability to use, existing tools and applications that facilitate parental control over children’s access to social media. In addition, parents were specifically asked about their familiarity with and ability to use the “Kids Wallet” application. Responses to both items were measured on a five-point Likert scale ranging from 1 (“strongly disagree”) to 5 (“strongly agree”). The “Kids Wallet” is a mobile application developed by the Greek government to support parents in managing their children’s digital safety, monitoring screen time, and verifying age for digital services. It operates as a digital identity tool for children, enabling parental supervision of online activity.

Specifically, the application allows parents to regulate access to applications and websites, as well as to control the duration of their children’s usage. Furthermore, it provides a secure mechanism for online age verification, helping to protect minors from inappropriate content and potential risks. The application also functions as a digital wallet where children can store their digital identity, thereby facilitating access to age-restricted services and platforms. Responses to the two items were summed up to produce a total score ranging from 2 to 10. This composite measure represented the variable “parental familiarity with digital parental control tools,” with higher scores indicating greater levels of familiarity.

### Ethical issues

Ethical approval for the study protocol was granted by the Ethics Committee of the Faculty of Nursing at the National and Kapodistrian University of Athens (Approval No. 71; April 21, 2026). The study was carried out in accordance with the principles outlined in the Declaration of Helsinki.^16^ Prior to participation, individuals were provided with comprehensive information about the study’s aims and procedures, and informed consent was obtained from all parents. Data collection was conducted anonymously, and participation was entirely voluntary.

### Statistical analysis

Categorical variables are reported as frequencies (n) and percentages (%), while continuous variables are summarized using descriptive statistics, including the mean, standard deviation (SD), median, minimum, and maximum values. The normality of continuous variables was evaluated using the Kolmogorov–Smirnov test in conjunction with Q–Q plots, both of which indicated a normal distribution. Sociodemographic characteristics, patterns of social media use, and levels of political engagement were treated as independent variables. The dependent variables comprised the scores for “confidence in the effectiveness of the social media ban”, “impact of the social media ban on children’s lives”, and “parental familiarity with digital parental control tools”. To assess the independent associations between the predictors and the outcome variables, multivariable linear regression analyses were performed. The results are presented as adjusted beta coefficients, along with their corresponding 95% confidence intervals (CIs), p-values, and adjusted R² values. Statistical significance was set at p < 0.05. All analyses were performed using IBM SPSS Statistics for Windows, version 28.0 (IBM Corp., Armonk, NY, USA).

## Results

### Sociodemographic characteristics

Study sample included 366 parents. In our sample, 57.1% were females. Mean age was 45.2 years, with a median value of 45, a minimum age of 28, and a maximum age of 63. Mean financial status score was 6.4, with a median score of 7, a minimum score of 3, and a maximum score of 10. Table 1 shows the sociodemographic characteristics of the parents.

**Table 1.**
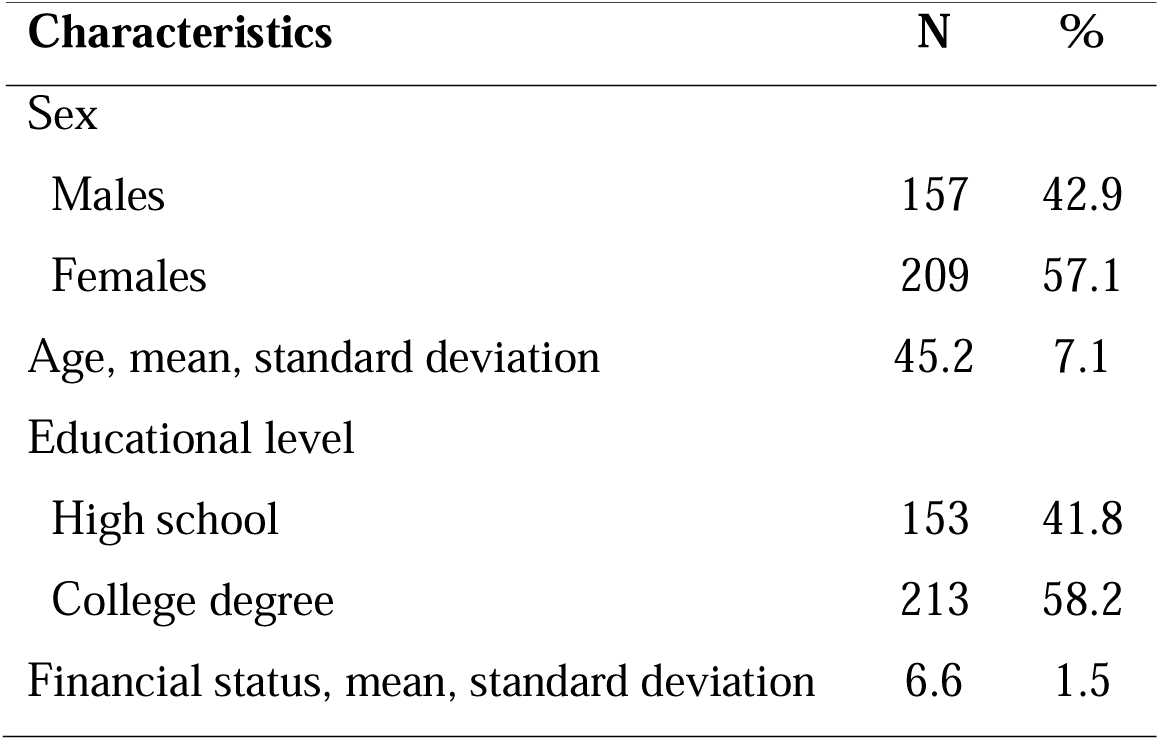
Sociodemographic characteristics of the parents.

### Social media use

Most parents had accounts on two platforms (36.9%), while 26.2% had accounts on one platform, 23.0% had accounts on three platforms, and 14.0% had accounts on 4-5 platforms. Mean daily time spent on social media was 2.0 hours, with a median time of 2 hours, a minimum value of thirty minutes, and a maximum value of 12 hours. In our sample, 43.4% did not post ever, 34.4% posted once a month on social media, 13.1% posted once in two weeks, 4.1% posted once a week, and 4.1% posted two-four days a week, and 0.8% posted five-seven days a week. Table 2 shows the social media use of parents.

**Table 2.** Social media use of the parents.

| **Social media use** | **N** | **%** |
| --- | --- | --- |
| Total number of accounts on social media |  |  |
| 1 | 96 | 26.2 |
| 2 | 135 | 36.9 |
| 3 | 84 | 23.0 |
| 4 | 39 | 10.7 |
| 5 | 12 | 3.3 |
| Daily time spent on social media, mean, standard deviation | 2.0 | 1.6 |
| Frequency of posting on social media |  |  |
| Never | 159 | 43.4 |
| About once a month | 126 | 34.4 |
| About once in two weeks | 48 | 13.1 |
| About once a week | 15 | 4.1 |
| Two-four days a week | 15 | 4.1 |
| Five-seven days a week | 3 | 0.8 |

### Political engagement

Among our parents, 78.4% reported that they follow somewhat/very closely news about politics, while 21.6% reported that they follow not closely at all/not very closely news about politics. Moreover, 45.9% reported that they talk sometimes to people about politics, 20.5% talk often/very often, and 33.6% talk rarely or not at all. Table 3 shows the political engagement of the parents.

**Table 3.** Political engagement of the parents.

| **Political engagement** | **N** | **%** |
| --- | --- | --- |
| How closely do you follow news about politics? |  |  |
| Not closely at all | 15 | 4.1 |
| Not very closely | 60 | 16.4 |
| Somewhat closely | 144 | 39.3 |
| Closely | 111 | 30.3 |
| Very closely | 36 | 9.8 |
| How often do you talk to people about politics? |  |  |
| Not at all | 33 | 9.0 |
| Rarely | 90 | 24.6 |
| Sometimes | 168 | 45.9 |
| Often | 63 | 17.2 |
| Very often | 12 | 3.3 |

### Opinion on social media ban for children

Table 4 shows parents’ opinion on social media ban for children. In the sample study, 54.1% of respondents reported that the appropriate age for a child to have a personal social media account is 17 years, followed by 24.6% who reported 16 years, 11.5% who reported 15 years, 4.1% who reported 14 years, 4.1% who reported 13 years, and 1.6% who reported 12 years. Most parents (84.4%) stated that parents should be responsible for setting limits on children’s use of social media, whereas 15.6% reported that this responsibility should rest with governments. Most parents stated that the problematic social media use among children constitutes an important public health concern (95.1%). In our sample, 68.0% agreed with the implementation of a social media ban for all children under age of 15. Also, most of our sample (91.8%) reported that they need more information from the government regarding the implementation of the ban. Additionally, 89.3% believed that additional measures, beyond a social media ban, should be implemented to address the problem.

Most parents agreed that children will be able to find ways to create accounts on platforms to which the social media ban will be applied (79.3%), and they will create accounts on platforms that will be free and, therefore, less regulated (85.3%). Also, only 36.1% believed that social media platforms will fully comply with the legislation regarding the social media ban.

In our sample, 51.6% reported that social media ban will improve children’s mental health and 55.8% reported that social media ban will improve children’s school performance (47.5%). Moreover, 64.8% reported that social media ban will improve children’s sleep quality.

### Parental familiarity with digital parental control tools

Among our parents, 27.9% stated that they are not familiar with and able to use available tools and applications that enable parental control over children’s access to social media. Additionally, 63.1% reported that they are not familiar with and able to use the application “Kids Wallet”.

In the sample, most parents favored targeted interventions over a social media ban, including digital literacy courses in schools (86.1%), active parental involvement in digital literacy (74.6%), prohibition of inappropriate content (77.9%), reasonable parental limits on social media use (73.8%), and restriction of addictive platform features (73.0%).

**Table 4.**
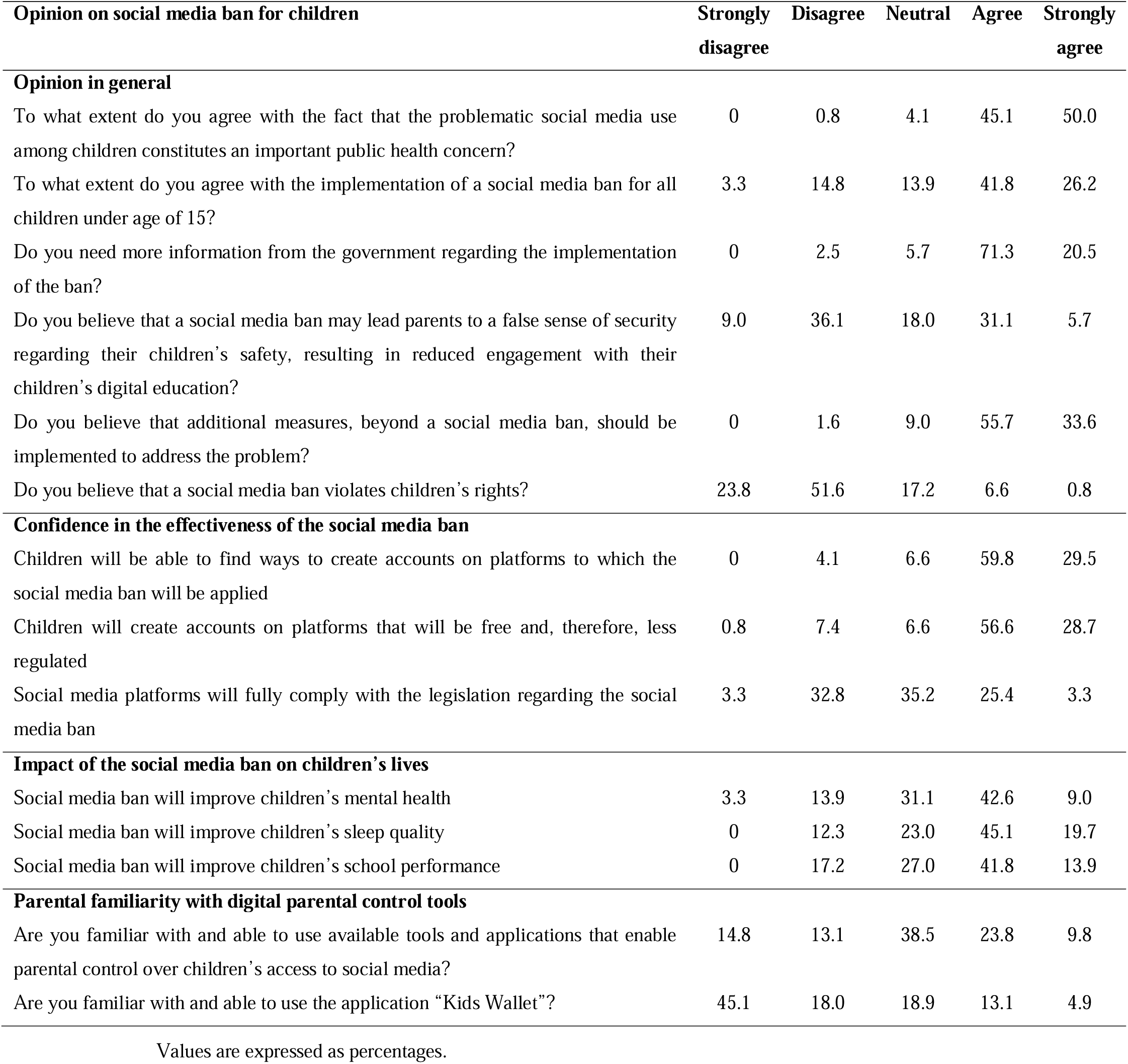
Parents’ opinion on social media ban for children.

| **Opinion on social media ban for children** | **Strongly disagree** | **Disagree** | **Neutral** | **Agree** | **Strongly agree** |
| --- | --- | --- | --- | --- | --- |
| **Opinion in general** |  |  |  |  |  |
| To what extent do you agree with the fact that the problematic social media use among children constitutes an important public health concern? | 0 | 0.8 | 4.1 | 45.1 | 50.0 |
| To what extent do you agree with the implementation of a social media ban for all children under age of 15? | 3.3 | 14.8 | 13.9 | 41.8 | 26.2 |
| Do you need more information from the government regarding the implementation of the ban? | 0 | 2.5 | 5.7 | 71.3 | 20.5 |
| Do you believe that a social media ban may lead parents to a false sense of security regarding their children's safety, resulting in reduced engagement with their children's digital education? | 9.0 | 36.1 | 18.0 | 31.1 | 5.7 |
| Do you believe that additional measures, beyond a social media ban, should be implemented to address the problem? | 0 | 1.6 | 9.0 | 55.7 | 33.6 |
| Do you believe that a social media ban violates children's rights? | 23.8 | 51.6 | 17.2 | 6.6 | 0.8 |
| **Confidence in the effectiveness of the social media ban** |  |  |  |  |  |
| Children will be able to find ways to create accounts on platforms to which the social media ban will be applied | 0 | 4.1 | 6.6 | 59.8 | 29.5 |
| Children will create accounts on platforms that will be free and, therefore, less regulated | 0.8 | 7.4 | 6.6 | 56.6 | 28.7 |
| Social media platforms will fully comply with the legislation regarding the social media ban | 3.3 | 32.8 | 35.2 | 25.4 | 3.3 |
| **Impact of the social media ban on children's lives** |  |  |  |  |  |
| Social media ban will improve children's mental health | 3.3 | 13.9 | 31.1 | 42.6 | 9.0 |
| Social media ban will improve children's sleep quality | 0 | 12.3 | 23.0 | 45.1 | 19.7 |
| Social media ban will improve children's school performance | 0 | 17.2 | 27.0 | 41.8 | 13.9 |
| **Parental familiarity with digital parental control tools** |  |  |  |  |  |
| Are you familiar with and able to use available tools and applications that enable parental control over children's access to social media? | 14.8 | 13.1 | 38.5 | 23.8 | 9.8 |
| Are you familiar with and able to use the application "Kids Wallet"? | 45.1 | 18.0 | 18.9 | 13.1 | 4.9 |

Values are expressed as percentages.

### Dependent variable: Confidence in the effectiveness of the social media ban

Table 5 reports the results of the multivariable linear regression model with score on the variable “confidence in the effectiveness of the social media ban” as the dependent variable. We found that increased age is associated with increased confidence in the effectiveness of the social media ban (adjusted coefficient beta = 0.072, 95% CI = 0.043 to 0.102, p<0.001). In other words, older parents showed higher levels of confidence in the effectiveness of the social media ban.

**Table 5.**
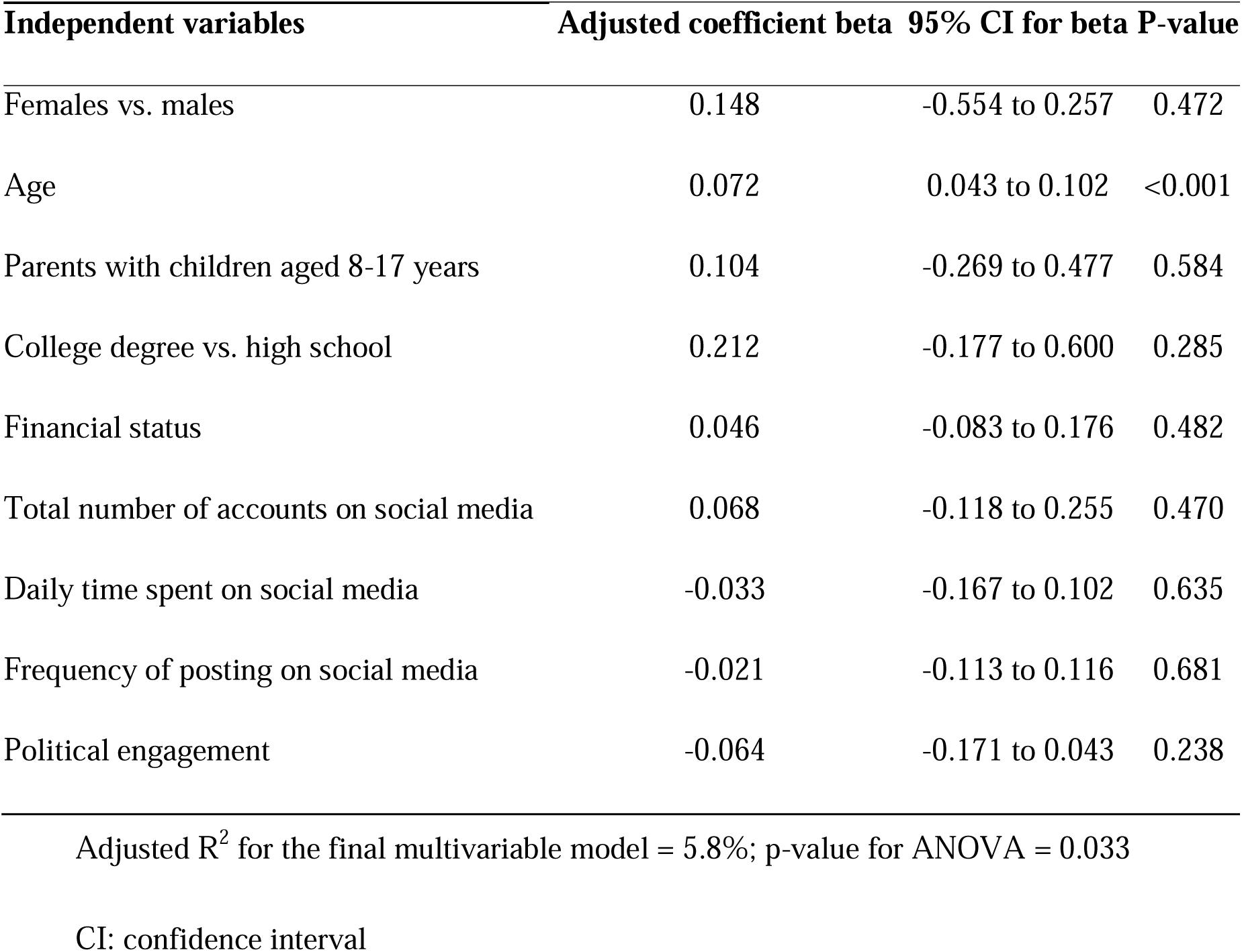
Multivariable linear regression model with score on the variable “confidence in the effectiveness of the social media ban” as the dependent variable.

| Independent variables | Adjusted coefficient beta | 95% CI for beta | P-value |
| --- | --- | --- | --- |
| Females vs. males | 0.148 | -0.554 to 0.257 | 0.472 |
| Age | 0.072 | 0.043 to 0.102 | <0.001 |
| Parents with children aged 8-17 years | 0.104 | -0.269 to 0.477 | 0.584 |
| College degree vs. high school | 0.212 | -0.177 to 0.600 | 0.285 |
| Financial status | 0.046 | -0.083 to 0.176 | 0.482 |
| Total number of accounts on social media | 0.068 | -0.118 to 0.255 | 0.470 |
| Daily time spent on social media | -0.033 | -0.167 to 0.102 | 0.635 |
| Frequency of posting on social media | -0.021 | -0.113 to 0.116 | 0.681 |
| Political engagement | -0.064 | -0.171 to 0.043 | 0.238 |

Adjusted $R^2$ for the final multivariable model = 5.8%; $p$ -value for ANOVA = 0.033
CI: confidence interval

### Dependent variable: Impact of the social media ban on children’s lives

Table 6 reports the results of the multivariable linear regression model with score on the variable “impact of the social media ban on children’s lives” as the dependent variable. We found that females (adjusted coefficient beta = 0.228, 95% CI = 0.134 to 0.320, p=0.012) and parents with a college degree (adjusted coefficient beta = 0.626, 95% CI = 0.105 to 1.148, p=0.019) have higher score on the variable “impact of the social media ban on children’s lives”. Moreover, our results showed a positive association between age (adjusted coefficient beta = 0.068, 95% CI = 0.041 to 0.092, p<0.001), financial status (adjusted coefficient beta = 0.253, 95% CI = 0.079 to 0.426, p=0.005), total number of accounts on social media (adjusted coefficient beta = 0.117, 95% CI = 0.043 to 0.213, p=0.033), daily time spent on social media (adjusted coefficient beta = 0.117, 95% CI = 0.043 to 0.213, p=0.013), and impact of the social media ban on children’s lives.

**Table 6.**
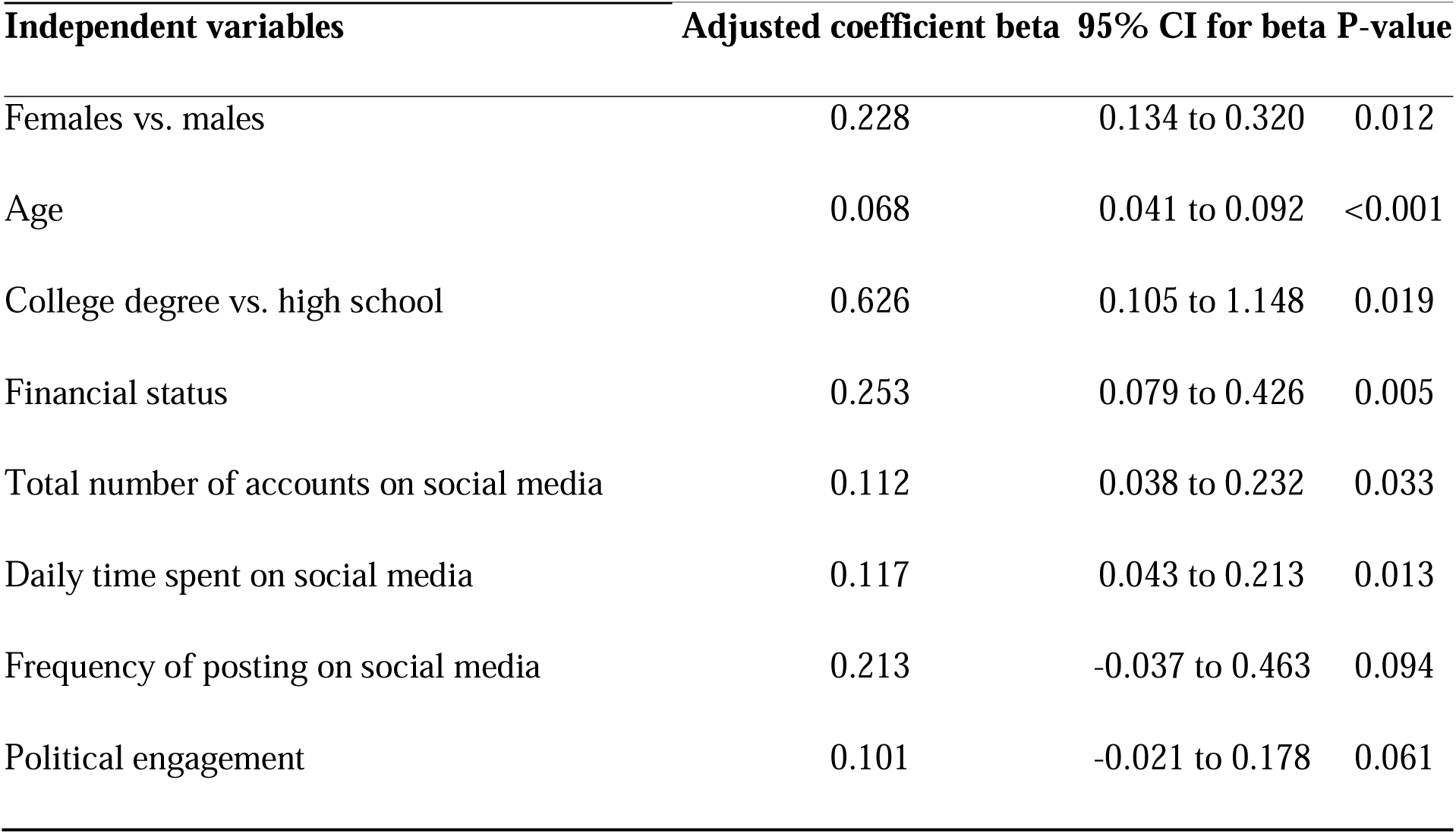
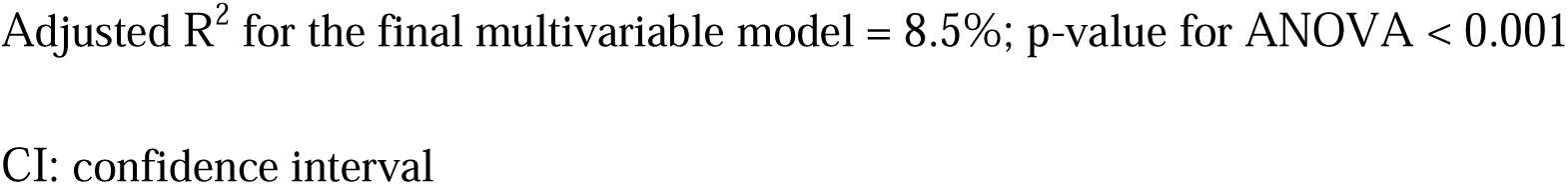
Multivariable linear regression model with score on the variable “impact of the social media ban on children’s lives” as the dependent variable.

| Independent variables | Adjusted coefficient beta | 95% CI for beta | P-value |
| --- | --- | --- | --- |
| Females vs. males | 0.228 | 0.134 to 0.320 | 0.012 |
| Age | 0.068 | 0.041 to 0.092 | <0.001 |
| College degree vs. high school | 0.626 | 0.105 to 1.148 | 0.019 |
| Financial status | 0.253 | 0.079 to 0.426 | 0.005 |
| Total number of accounts on social media | 0.112 | 0.038 to 0.232 | 0.033 |
| Daily time spent on social media | 0.117 | 0.043 to 0.213 | 0.013 |
| Frequency of posting on social media | 0.213 | -0.037 to 0.463 | 0.094 |
| Political engagement | 0.101 | -0.021 to 0.178 | 0.061 |

Adjusted $R^2$ for the final multivariable model = 8.5%; p-value for ANOVA < 0.001
CI: confidence interval

### Dependent variable: Parental familiarity with digital parental control tools

Table 7 reports the results of the multivariable linear regression model with score on the variable “parental familiarity with digital parental control tools” as the dependent variable. We found that reduced age is associated with increased parental familiarity with digital parental control tools (adjusted coefficient beta = -0.054, 95% CI = -0.121 to -0.008, p=0.037). Additionally, multivariable analysis identified a positive association between total number of accounts on social media (adjusted coefficient beta = 0.283, 95% CI = 0.066 to 0.499, p=0.011) and parental familiarity with digital parental control tools.

**Table 7.** Multivariable linear regression model with score on the variable “parental familiarity with digital parental control tools” as the dependent variable.

| Independent variables | Adjusted coefficient beta | 95% CI for beta | P-value |
| --- | --- | --- | --- |
| Females vs. males | 0.279 | -0.192 to 0.750 | 0.245 |
| Age | -0.054 | -0.121 to -0.008 | 0.037 |
| College degree vs. high school | -0.334 | -0.785 to 0.118 | 0.147 |
| Financial status | 0.155 | -0.014 to 0.512 | 0.064 |
| Total number of accounts on social media | 0.283 | 0.066 to 0.499 | 0.011 |
| Daily time spent on social media | -0.004 | -0.161 to 0.153 | 0.960 |
| Frequency of posting on social media | 0.081 | -0.135 to 0.298 | 0.460 |
| Political engagement | 0.111 | -0.022 to 0.321 | 0.114 |

Adjusted R<sup>2</sup> for the final multivariable model = 8.3%; p-value for ANOVA < 0.001
CI: confidence interval

## Discussion

Following Australia’s implementation of a nationwide ban on social media access for individuals under the age of 16 in December 2025, a number of countries have introduced legislation mandating age verification on social media platforms to reduce online risks. For example, several European nations are currently exploring comparable measures, while the European Commission has indicated its commitment to supporting a coordinated, region-wide strategy through the recent completion of a standardized age verification system. Within this evolving regulatory landscape, assessing public attitudes toward banning social media use among children is crucial, as societal perspectives significantly influence the acceptability, legitimacy, and overall effectiveness of such policy interventions. To the best of our knowledge this is the first study that examines parents’ opinion toward social media ban for children and its influencing factors.

A central finding of this study is that a very large proportion of parents (95.1%) identified problematic social media use among children as a significant public health issue. This strong consensus indicates that worries about children’s interactions with social media extend beyond individual households and are increasingly viewed as a broader societal concern requiring population-level responses. Recognizing problematic social media use as a public health matter is essential for fostering support for and ensuring the success of regulatory and preventive strategies. In line with this perspective, our results are consistent with a growing body of research documenting the adverse effects of excessive or problematic social media engagement on children’s mental health, overall wellbeing, and developmental outcomes.^17–21^ The widespread perception of risk observed in our sample may help explain the growing public acceptance of age-related restrictions and regulatory initiatives seen across different countries. Thus, the near-unanimous acknowledgment of this issue highlights the importance of evaluating not only the effectiveness but also the social acceptability of interventions such as restricting children’s access to social media platforms.

Moreover, a substantial majority of parents (91.8%) indicated that they require clearer and more comprehensive information from governmental authorities regarding how a social media ban would be implemented. Evidence from Australian parents supports this finding, with 47% reporting that they do not understand the functioning of the government’s social media ban policy. There is also a clear need to strengthen parental education on digital control tools, as only 27.9% of parents in our sample reported familiarity with such resources. Awareness is even lower for the “Kids Wallet” application—a Greek government mobile tool designed to help parents oversee their children’s online safety—with just 18.0% indicating familiarity. Taken together, these results reveal a significant gap between general support for regulatory measures and understanding of their practical implementation. The strong demand for official guidance suggests that public acceptance of a social media ban depends heavily on clear communication regarding its scope, enforcement procedures, age verification methods, and the respective responsibilities of parents, schools, and platform providers. Inadequate communication may lead to confusion, mistrust, or unrealistic expectations about the protective impact of such policies. From a public health standpoint, transparent and proactive information dissemination is crucial not only to promote compliance but also to avoid fostering a false sense of security that could diminish parental involvement in children’s digital education.^22–24^

This study identifies a positive relationship between age and confidence in the effectiveness of a social media ban, as well as between age and perceptions of its beneficial impact on children’s lives. These findings offer valuable insight into generational variations in attitudes toward regulatory measures. Older parents may be more likely to regard legislative interventions as effective means of managing risk, potentially reflecting greater trust in institutional authority or more familiarity with public health policies shaped by regulation. Indeed, older individuals tend to exhibit higher levels of trust in public institutions compared to younger, more skeptical cohorts, and this trust often translates into stronger support for governmental action. In contrast, younger respondents—who are typically more immersed in digital environments—may view such restrictions as less practical or enforceable.^25,26^ Furthermore, increasing age is often linked to a heightened awareness of long-term societal and health implications, which may reinforce beliefs in the preventive value of regulatory approaches.^27^ Differences in risk perception may also contribute, as older individuals are more inclined to prioritize long-term developmental and societal outcomes, whereas younger people may focus more on immediate personal experiences.^28^ Overall, these findings suggest that confidence in the effectiveness of social media restrictions is influenced not only by perceived digital risks for children but also by broader life experience, normative views on governance, and attitudes toward state intervention. Recognizing these age-related differences is crucial for anticipating public reactions to policy implementation and for designing communication strategies that effectively address skepticism among younger groups, who may have greater exposure to social media and stronger reservations about the feasibility of such measures.

Another key finding of this study is that parents with higher socioeconomic status—reflected in both educational attainment and financial standing—were more likely to view a social media ban as beneficial for children. Higher socioeconomic status has been linked to greater exposure to scientific knowledge, enhanced health literacy, and a stronger ability to assess complex risk–benefit considerations, particularly within public health contexts.^29,30^ As a result, these individuals may be more informed about evidence highlighting the potential harms of excessive or problematic social media use among children and, consequently, more supportive of preventive, population-level interventions. Furthermore, higher levels of education are often associated with stronger endorsement of evidence-based policymaking and increased trust in regulatory institutions, especially when policies aim to protect collective interests such as child wellbeing. By contrast, individuals with lower educational levels are more likely to express skepticism toward regulatory measures, frequently due to concerns about their feasibility, possible unintended consequences, or perceived intrusion into private and family life.^31^

An additional finding of this study is that parents who reported higher levels of social media use—such as spending more time on these platforms daily—were more likely to perceive a social media ban as beneficial for children. This pattern may point to the influence of personal experience in shaping both risk perception and attitudes toward regulation. Individuals who use social media extensively are more likely to encounter its negative aspects, including compulsive usage patterns, displacement of time from other activities, emotional strain, and exposure to potentially harmful or distressing content.^32,33^ Such direct exposure may encourage a more critical and reflective view of social media environments, particularly regarding their appropriateness for children, whose self-regulatory abilities are still developing. Frequent users may also be more aware of persuasive design elements—such as infinite scrolling, algorithm-driven content personalization, and continuous notifications—intended to maximize user engagement.^34^ This awareness can increase support for external regulatory approaches, including bans, as protective measures for younger users who may be more susceptible to these influences. In contrast, individuals with lower levels of social media engagement may have less direct experience with these risks and challenges, and therefore may be less inclined to view restrictive measures as necessary or beneficial.

A related finding in our study is that parents who reported higher levels of social media engagement—reflected in a greater number of social media accounts—also demonstrated greater familiarity with digital parental control tools. Parents’ digital competencies and their active involvement in online environments play a crucial role in their ability to implement effective supervision strategies, including the use of monitoring technologies and technical safeguards.^35^ More frequent interaction with social media platforms may enhance technical skills and increase awareness of platform-specific safety features, thereby strengthening parents’ confidence in using parental control tools. Digitally engaged parents are therefore more likely to adopt technological solutions rather than relying exclusively on restrictive or avoidance-based approaches. Additionally, a deeper understanding of social media ecosystems may heighten awareness of potential online risks, encouraging more proactive efforts to protect children through the use of available digital tools.^36,37^ Conversely, parents with lower levels of social media use may encounter structural and knowledge-related barriers that limit both their understanding and adoption of parental control technologies, even when such tools are accessible.

Several limitations should be taken into account when interpreting the results of this study. First, the cross-sectional design limits the ability to establish causal relationships between parents’ characteristics and their attitudes toward a social media ban. Second, the reliance on self-reported data introduces the possibility of reporting biases, such as social desirability and recall bias. Third, although efforts were made to include a diverse range of sociodemographic variables, the sample may not fully represent the broader population. Fourth, attitudes toward a social media ban were examined within a hypothetical or prospective policy framework. Public perceptions may shift once such measures are implemented, particularly as individuals gain direct experience with enforcement procedures, age verification systems, and potential unintended effects. Fifth, despite examining multiple relevant factors, some important variables may not have been captured. Finally, the rapidly changing landscape of digital technologies and social media platforms constitutes an inherent limitation, as both public attitudes and policy preferences may evolve in response to new platform features, regulatory developments, or emerging evidence on the effects of social media on children’s wellbeing. Despite these limitations, the study offers important insights into public views on social media restrictions for children and identifies key considerations for policymakers. Future research using longitudinal designs, more representative samples, and qualitative methodologies could provide a deeper understanding of how public attitudes evolve and how they interact with policy implementation over time.

In conclusion, this study explored parents’ attitudes toward a potential social media ban for children and identified key factors shaping their views on this policy measure. The results reveal a high level of societal concern about children’s engagement with social media, with most parents recognizing problematic use as a major public health issue. This broad acknowledgment positions children’s digital wellbeing firmly within the public health domain and supports the case for proactive governmental action.

Importantly, although a considerable proportion of respondents expressed support for implementing a social media ban, this support was not unconditional. Parents consistently highlighted that restrictive policies should not function in isolation but rather be incorporated into a wider, integrated policy framework. The strong endorsement of complementary measures reflects a preference for a comprehensive, preventive approach rather than one that relies solely on prohibition.

From a policy standpoint, these findings carry several implications. First, social media restrictions for children are more likely to gain social acceptance and encourage compliance when they are presented as part of a multi-faceted strategy that combines protection with empowerment. Policymakers should therefore avoid framing bans as standalone interventions and instead clarify how such measures will operate alongside educational initiatives, family-level responsibilities, and platform-based regulations. Second, the observed variations in attitudes across sociodemographic groups and levels of social media use highlight the importance of targeted communication strategies. Clear, accessible, and tailored messaging will be essential to address skepticism, manage expectations, and promote inclusive engagement across different population groups.

Overall, the findings suggest that effective regulation of children’s social media use requires moving beyond simplistic, binary debates about bans toward more holistic, evidence-based policy approaches. Such frameworks should integrate regulation, education, and shared responsibility. Policymakers are encouraged to build on existing public support for child protection while also investing in digital literacy, strengthening parental capacity, and enhancing platform accountability to achieve sustainable and equitable outcomes.

## Data Availability

All data produced in the present study are available upon reasonable request to the authors

